# Associations between adult obesity and mid-life weight change patterns with cardiometabolic health in early old age: Evidence from the 1958 British birth cohort

**DOI:** 10.64898/2026.08.26.26361387

**Authors:** Charis Bridger Staatz, Laura Gimeno, Naveed Sattar, Nish Chaturvedi, George Ploubidis

## Abstract

**Background:** Cardiometabolic health typically declines with age and is worse among individuals living with obesity. Weight loss medications have modified the potential for weight loss across the life course, but it remains unclear whether weight reduction in later midlife contributes to improved cardiometabolic health, or if continuing to gain weight may continue to worsen cardiometabolic health.

**Methods:** Using the nationally representative 1958 National Child Development Study (NCDS), a British birth cohort, associations were examined using lagged linear regression between weight change between ages 50-55 and health outcomes at age 62 (n=6,309 high-density lipoprotein (HDLc) and low-density lipoprotein (LDLc) cholesterol, systolic and diastolic blood pressure (SBP and DBP), heart rate, triglycerides, C-reactive protein (CRP), and glycated haemoglobin (HbA1c). Models accounted for prior biomarker levels at age 44. We also explored impacts of weight change on subsequent body composition.

**Results:** Those who gained weight into or within obesity had less favourable cardiometabolic profiles and experienced faster deterioration of cardiometabolic markers between the ages of 44 and 62 than those remaining in healthy weight (e.g. SBP: 5.726, 95% CI: 2.660 to 8.793, p < 0.001; CRP: 0.802, 95% CI: 0.409 to 1.196, p < 0.001). Those who lost weight from obesity had similar *rates* of cardiometabolic biomarker deterioration to the healthy weight group (SBP: 0.947, 95%CI: -6.605 to 8.499, p=0.806; CRP: 0.140, 95% CI: -0.774 to 1.055, p= 0.764).

**Conclusion:** Weight change in midlife tends towards increasing obesity and associated adverse cardiometabolic risk. Those who lose weight experienced improved cardiometabolic profiles. By viewing midlife as a modifiable stage of the life course, this study highlights opportunities to promote cardiometabolic health, and limit the speed of health decline.

**Funding:** BRC UCLH, ESRC

## Introduction

Obesity has consistently and robustly been linked to increased morbidity and all-cause mortality [1], including to an increased risk of cardiovascular disease and cancer [2]. Duration of exposure to obesity is also an important cardiovascular disease risk factor [3], and while evidence suggests that excess weight gain before age 30 may be particularly harmful for some aspects of health [4], the impact of chronic obesity exposure remains poorly understood.

Addressing this gap is important because levels of obesity in the UK are rising fastest among young adults [5], and obesity has become more prevalent and is developing earlier in more recently born cohorts [6, 7], a trend that is becoming known as a “generational health drift” [8]. As cohort life expectancy continues to increase and obesity onset shifts to younger ages, average time exposed to obesity over the life course is increasing.

Obesity management medications (OMM; based on incretin therapies) have the potential to support effective weight loss across the population. In the UK at the start of 2025, approximately 3% of people were using OMM with greatest use concentrated among adults aged 45-55 years, and a further 6.5% stated interest in using them in the next year [9]. Evidence from the British birth cohorts suggests that weight loss was uncommon in the pre-OMM era, whilst continued weight gain in later midlife is widespread, affecting over one-third of adults already living with obesity in their early forties [7].

Different weight change patterns across life have the potential to differently shape cardiovascular risk in older age, as well as how cardiovascular risk *changes* as individuals age. Given that more recent generations are already experiencing earlier obesity onset and the opportunity that OMMs present for modifying weight trajectories, it is necessary to establish whether successful weight loss in later life can improve cardiometabolic outcomes compared to alternative scenarios for those living with obesity. This includes understanding the consequences of continued weight gain into later mid-life. Understanding the health implications of these contrasting weight trajectories can help us to anticipate the long-term benefits of supporting medicated weight loss, and of preventing further weight gain among those at greatest risk. By viewing health through the lens of change, rather than a status at a single point, we can also identify factors that may help limit the speed of health decline.

Understanding modifiable influences on the rate of cardiometabolic health deterioration, particularly during midlife – a period increasingly recognised as important for shaping aging trajectories [10] - is central to promoting cardiometabolic health in early old age. Using rich life course data from the nationally representative 1958 National Child Development Study (1958) in Britain, we examined how weight change in people’s 50s was associated with the rate of change in cardiometabolic biomarkers by the time they reached their 60s.

## Methods

### Data

The 1958 British National Child Development Study (NCDS) is a longitudinal cohort study that follows the lives of approximately 17,500 individuals born in England, Scotland and Wales during a single week in 1958 [11]. Participants have been followed up on 10 occasions since birth and data on social, biological and physical health characteristics have been prospectively collected. When respondents were aged 44-45 (2002), 9377 cohort members responded to a biomedical survey. Another biomedical survey occurred at ages 61-65 (2020-24), with 6,309 respondents, representing the analytic sample.

### Outcomes

At the biomedical sweeps, biomarkers indicative of cardiometabolic risk were measured from non-fasting blood samples, including total cholesterol, high-density lipoprotein cholesterol (HDLc), triglycerides, C-reactive protein (CRP) and HbA1c (glycated haemoglobin). Using information on total cholesterol, HDLc and triglycerides, we derived low-density lipoprotein cholesterol (LDLc). Mean systolic and diastolic blood pressure were calculated from two seated readings, and heart rate was measured at the same time. All biomarker measures are reported as continuous measures, and corrections for medication use have been applied. At age 62, body fat percentage (BF%) was also collected for the first time, from which fat mass (FM), fat free mass (FFM) and fat mass index (FMI) were calculated, and the ratio of FM and FFM was derived (FM/FFM). See Table S1 for further description of these measures.

### Exposure

At each data collection sweep in adulthood for which height and weight data were available, we derived BMI. To identify later-life weight loss, peak BMI was defined as the highest observed BMI between ages 23 and 42. Sustained weight loss occurring between ages 50 and 55 was defined as:

*More than 5% of body weight lost from a previous peak BMI greater than 30 kg/m^2^ (obesity) or 25 kg/m^2^ (overweight) and sustained for at least two time points spanning an average of 5 years*.

We split the weight loss groups for individuals whose peak BMI was greater than 30 kg/m^2^ (obesity) and between 25kg/m^2^ and 30 kg/m^2^ (overweight). We also identified individuals who maintained a healthy weight, overweight or obesity during the observation window; individuals who gained >5% of body weight within or into overweight or obesity; and those who weight cycled in and out, or within obesity during the observation window. Further details on this classification are provided in the Supplementary Material S2.

### Covariates

Covariates from early life and early adulthood up to age 23 were included in the analysis. This cut-off corresponds to the earliest BMI measurement contributing to peak BMI, ensuring that all covariates were measured before the earliest point at which the weight change could occur.

Early life measures were birthweight and parental occupational social class at birth. Cognitive ability was assessed at age 11. BMI and psychological distress (high/low malaise) were measured at age 16. Adult covariates measured at age 23 included self-rated health (excellent / good / fair or poor), smoking status (never smoked / ex-smoker / current smoker), occupational social class (manual or non-manual), partnership status (in partnership or not), voting behaviour (voted in last election or not), trade union membership (yes / no), educational attainment (any post-school qualification), family weekly income, and membership in civic or social organisations (yes / no).

### Statistical Analysis

We first describe mean BMI across adulthood by weight change group, mean levels of each biomarker at ages 44 and 62, and measures of body composition at age 62. We then plot the mean BMI and mean values for all biomarkers at age 62, stratified by weight change group membership.

We examined associations between weight change groups at ages 50–55 and biomarker levels at age 62 using lagged linear regression models using imputed data combined with non-response weights to account for item nonresponse and attrition [12–15]. Model 1 adjusted for age at outcome measurement and sex. Model 2 additionally adjusted for the corresponding biomarker measured at age 44. Model 3 additionally adjusted for a range of potential confounders measured up to and including age 23 (described above), corresponding to the earliest point at which BMI contributing to the exposure definition was observed. Model 3 also adjusted for the age at which peak BMI was reached between ages 23 and 42, and for all other biomarker measures at age 44.

To understand the mechanistic relationship between weight change and subsequent body composition, which may mediate the relationship between weight change and biomarkers at age 62, we also explored associations between weight change groups and FMI and FM:FFM at age 62. These analyses show how weight change between age 50-55 relates to body composition at age 62, rather than to *change* in body composition between 44 and 62, as is the case for other biomarkers, since equivalent FMI, FM and FFM measures were not available at age 44. Model 1 adjusts for age at outcome measurement and sex only. Model 2 adjusts for covariates measured up to and including age 23 listed above, and age of peak BMI.

To assess the extent to which the observed associations may reflect causal effects, specifically the adequacy of the adjustment set and the potential impact of residual confounding, we used two negative control outcomes that would in theory be associated with the weight-change exposure only through shared confounders (e.g. socioeconomic position) [16]. These two controls were: financial literacy (defined as answering three questions correctly) and receipt of inheritance (categorised as none/one, or two or more inheritances). More information can be found in the Supplementary Material.

### Missing Data and Multiple Imputation

To address item missingness, we imputed missing data among respondents to the biomedical sweep at age 62 using multiple imputation with chained equations (MICE) in Stata. To increase the plausibility of the Missing at Random (MAR) assumption, imputation models included auxiliary variables predictive of both missingness and missing values (see Supplementary Material). These include a wide range of early-life, socioeconomic, behavioural, and health indicators. A total of 20 imputed datasets were generated, and estimates across imputations were combined using Rubin’s rules. To address the impact of attrition, we also used a non-response weight developed for the biomedical sweep at age 62 [12–15].

## Results

Among all participants assessed at age 62, only 1.85% had lost weight from obesity and 3.77% from overweight between the ages of 50 and 55 (Table 1). Approximately 24.4% maintained a healthy weight into their early 50s. The majority comprised individuals who maintained overweight (20.85%), gained weight into or within overweight (18.31%) or obesity (18.33%), experienced obesity weight cycling (7.58%), or maintained obesity without substantial weight change (4.91%).

**Table 1.** Prevalence of weight change groups at age 50 to 55.

| Weight Change Groups 50-55 | Estimated prevalence<br>(95% CI) |
| --- | --- |
| Always maintains normal weight | 24.40 (22.30 to 26.50) |
| Mostly maintains overweight | 21.07 (19.14 to 23.00) |
| Weight gain (>5%) - final weight overweight | 18.31 (16.38 to 20.24) |
| Weight loss (>5%) - peak weight overweight | 3.84 (2.35 to 5.33) |
| Mostly maintains Obese | 4.86 (3.71 to 6.02) |
| Weight gain (>5%) - final weight obese | 18.41 (16.39 to 20.42) |
| Weight loss (>5%) - peak weight obese | 1.80 (1.18 to 2.43) |
| Weight cycling - obese | 7.31 (5.91 to 8.72) |

Individuals who lost weight from obesity in their early–mid 50s typically had the highest mean BMI between ages 16 and 42 (Table S4). In contrast, those who gained weight into obesity generally had lower BMI in adolescence and early adulthood compared to the other obesity groups but saw more rapid weight gain between ages 42 and 55, resulting in the highest mean BMI by age 62. A similar pattern was observed among the overweight groups.

### Descriptive biomarker and body composition patterns

Mean biomarker levels at ages 44 and 62 by weight change groups at age 50-55 are shown in Table 2 and plotted against mean BMI at age 62 in Figure 1. At ages 44 and 62, individuals who maintained a healthy weight had the most favourable cardiometabolic profiles. Those who maintained overweight or were on trajectories toward overweight had intermediate profiles, whilst all those in the obesity range had the most adverse profiles.

**Figure 1.**
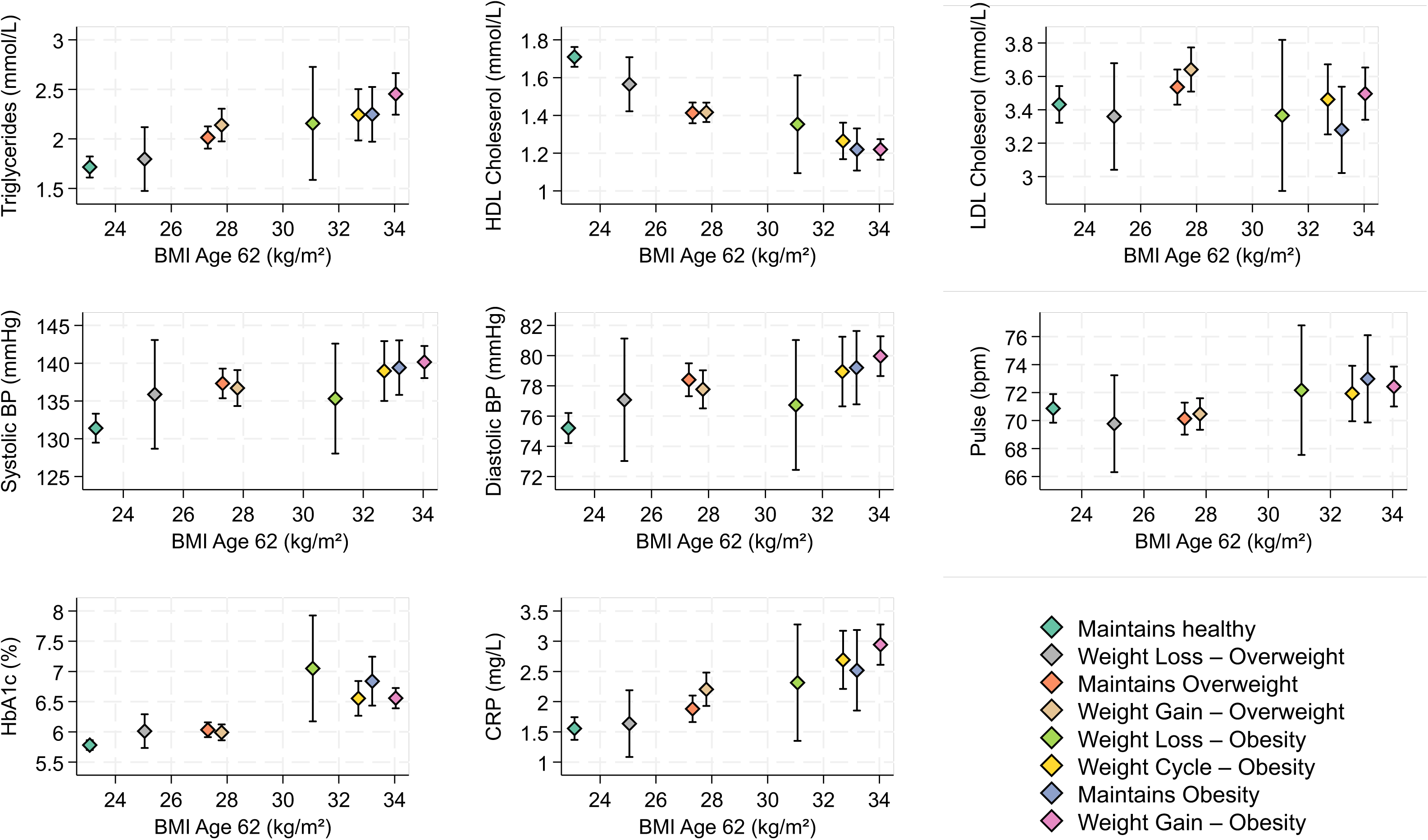
Mean estimated biomarker values at age 62 and weight change groups (ages of 50-55). ***Footnote.*** *Mean biomarker levels plotted against mean body mass index (BMI) at age C2, stratified by weight change groups at age 50-55. All biomarkers were analysed on their original scale. Acronyms are: Blood Pressure (BP), C-Reactive Protein (CRP), High Density Lipoprotein (HDL) Cholesterol; Low Density Lipoprotein (LDL) Cholesterol*.

**Figure 2.**
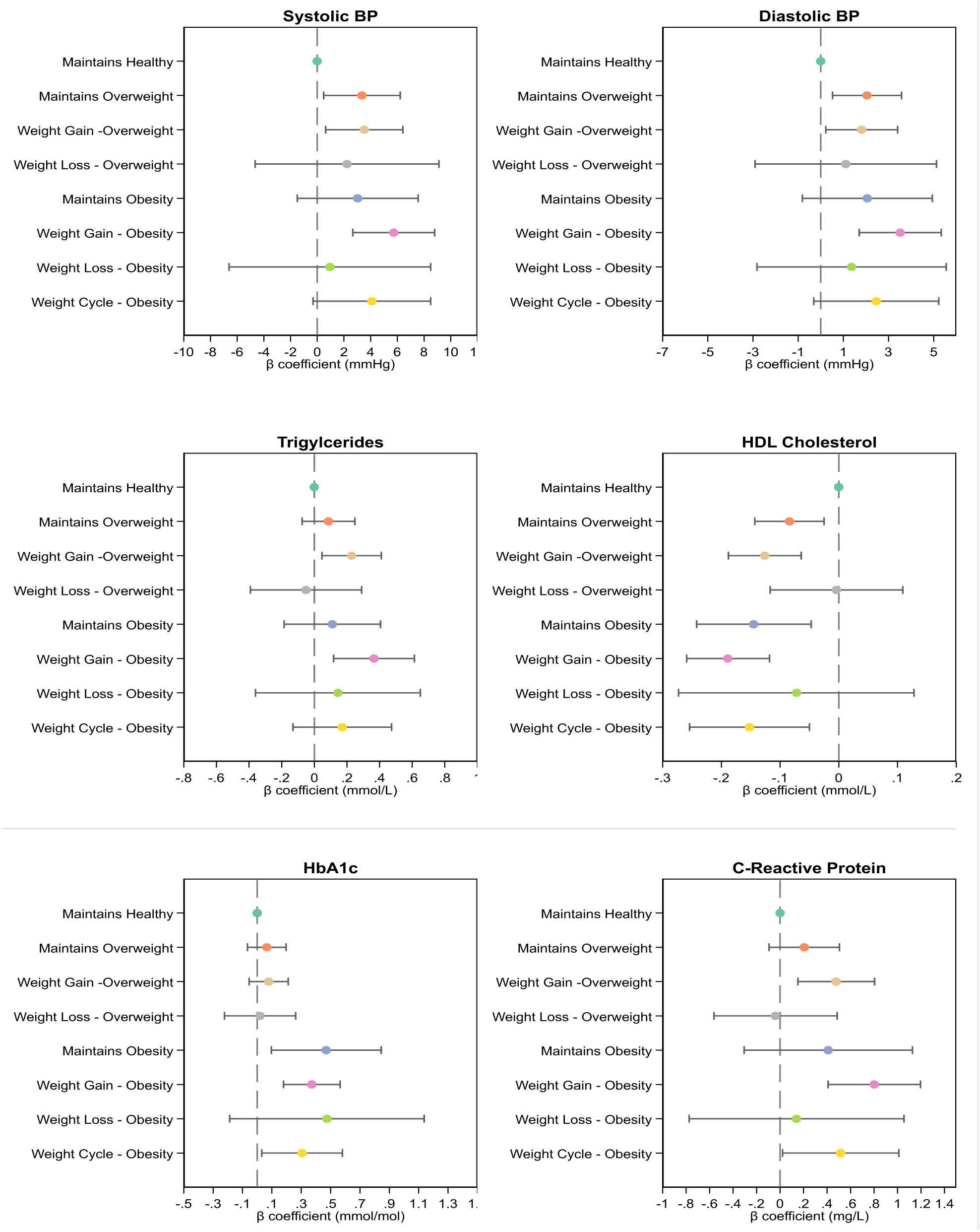
Fully adjusted Relative Risk Ratio (RRR) from linear lagged models for association between weight change and maintenance group and selected lagged biomarkers. *Footnote. Results from Model 3 for lagged linear regression testing association between weight change at age 50- 55 and rate of change in selected biomarkers between the age of 44 and C2 (full results for all lagged biomarkers available in Table S5). Models are adjusted for age at outcome measurement and sex, birthweight, parental occupational social class at birth, cognitive ability at age 11, BMI and psychological distress at age 1C, adult covariates measured at age 23 (self-rated health, smoking status, occupational social class, marital status, voting behaviour, trade union membership, educational attainment, income, and membership in civic or social organisations), age peak weight was achieved up to and including age 42, and all other biomarker measures at age 44, including the corresponding measures to the outcome*.

**Table 2.**
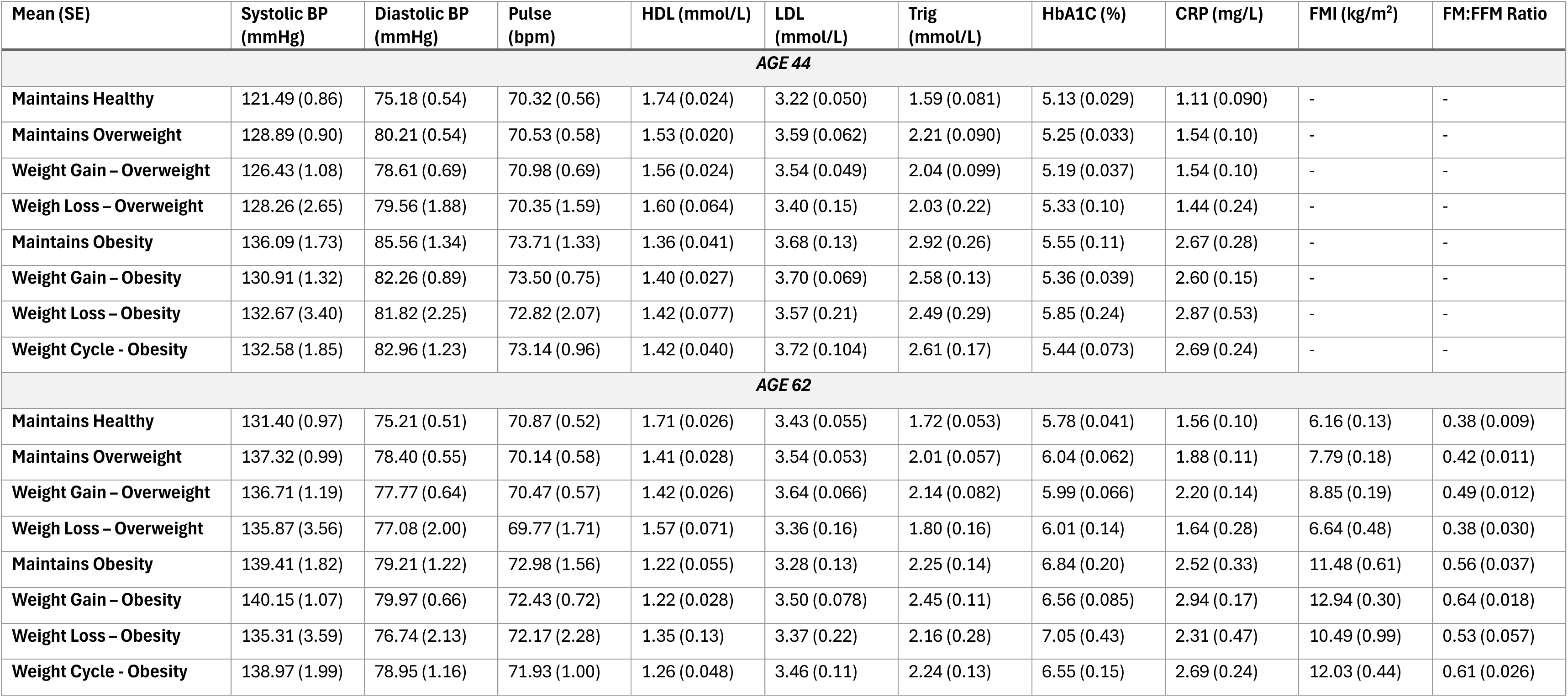
Mean values of blood biomarkers, blood pressure and body composition at ages 44 and 62, by weight change groups at age 50-55.

Descriptively, individuals who lost weight from overweight had mean biomarker levels at age 62 that more closely resembled those of the consistently healthy weight group. In contrast, those who lost weight from obesity had mean biomarker levels at age 62 that appeared more similar to those in the healthy weight group than was the case for other obesity-related groups, particularly the weight gain group. For HbA1c, mean levels at age 62 were elevated or similar in those who lost weight from obesity compared to the other obesity related groups.

Figure 1 plots estimated mean levels of each biomarker against BMI at age 62 according to weight change group membership between the ages of 50-55. Participants who maintained a healthy weight up to their mid-50s had the lowest BMI at age 62 (23.08 [95%CI: 22.80 to 23.36]), followed by those who lost weight from overweight (25.05 [95%CI: 24.07 to 26.02]), maintained overweight (27.31 [95%CI: 26.94 to 27.68]), and gained weight within the overweight range (27.80 [95%CI: 27.44 to 28.17]). Among obesity-related trajectories, those who lost weight from obesity during later midlife had the lowest BMI at age 62 (31.07 [95%CI: 29.28 to 32.86]), followed by weight cycling (32.70 [95%CI: 31.81 to 33.60]) and obesity maintenance (33.20 [95%CI: 32.18 to 34.22]). Those who gained weight into or within obesity in their early 50s had the highest BMI at age 62 (34.04 [95%CI: 33.53 to 34.55]).

Across the weight trajectory groups, a higher mean BMI at age 62 was generally associated with higher mean levels of triglycerides, systolic and diastolic blood pressure, and CRP, lower HDLc, and to a lesser extent higher heart rate, demonstrating largely linear relationships between BMI and each respective biomarker. HbA1c showed a threshold pattern, with higher mean levels observed across all obesity-related groups - whose BMI at age 62 remained greater than 30kg/m^2^ - while values were lower and similar among the overweight and healthy weight groups, despite differences in BMI at age 62. In contrast, there was no clear relationship between LDLc at age 62 across the different weight trajectory groups.

### Associations between weight change and maintenance groups and biomarker outcomes

Associations between weight change at ages 50–55 and biomarkers at age 62 in age- and sex-adjusted models mirrored the descriptive patterns (Table S5). However, adjustment for lagged biomarkers, covariates, and age of peak BMI showed that for most outcomes, those who lost weight from either overweight or obesity fared similarly to the healthy weight group in terms of the *rate* of biomarker deterioration between the ages of 44 and 62.

For systolic blood pressure, those who lost weight from either overweight or obesity showed no evidence of additional increase compared with the healthy weight group (weight loss– overweight: β: 2.234,95%: -4.655 to 9.124, p= 0.525; weight loss– obesity: β: 0.947,95%: -6.605 to 8.499, p= 0.806). In contrast, weight gain was associated with substantially greater increases, particularly for those gaining into or within obesity (weight gain–overweight: β: 3.522, 95%: 0.625 to 6.418, p= 0.017; weight gain–obesity β: 5.726,95%: 2.660 to 8.793, p < 0.001), and for those who weight cycled (β: 4.092, 95%: -0.318 to 8.502, p= 0.069). A similar pattern was observed for diastolic blood pressure.

For HDLc, those who lost weight from either overweight or obesity showed changes comparable to the healthy weight group (weight loss–overweight: β: -0.004, 95%: -0.117 to 0.109, p= 0.946; weight loss–obesity: β: -0.072, 95%: -0.273 to 0.128, p= 0.479). In contrast, weight gain was associated with declines in HDLc, particularly for those gaining into or within obesity (weight gain–overweight: β: -0.126, 95%: -0.188 to -0.064, p < 0.001; weight gain–obesity: β: -0.189, 95%: -0.259 to -0.118, p < 0.001). Those who weight cycled within/into obesity also experienced a greater decline in HDLc (β: -0.152, 95%: -0.254 to -0.050, p= 0.003). For LDLc and heart rate, there was little evidence of meaningful differences in rate of change between groups. For triglycerides, those who gained within or into overweight (β: 0.229, 95%: 0.047 to 0.411, p= 0.013) and obesity (β: 0.366, 95%: 0.118 to 0.614, p= 0.004) experienced greater increases compared to the healthy weight group. Those who lost weight had similar change in triglycerides as the healthy weight group (weight loss–overweight: β: -0.051, 95%: -0.392 to 0.290, p= 0.77; weight loss–obesity: β: 0.145, 95%: -0.361 to 0.650, p= 0.575).

For HbA1c, all groups with a history of obesity showed a higher rates of deterioration than the healthy weight group (obesity maintenance: β: 0.470, 95%: 0.096 to 0.845, p= 0.014; weight gain - obesity β: 0.372, 95%: 0.179 to 0.565, p< 0.001; obesity weight cycling β: 0.306, 95%: 0.031 to 0.580, p= 0.029), although for weight loss from obesity the effect was elevated but not significant (β: 0.475, 95%: -0.187 to 1.137, p= 0.16).

CRP showed a broadly similar pattern to systolic and diastolic blood pressure and triglycerides, with faster deterioration among those who gained weight into overweight (β: 0.478, 95%: 0.152 to 0.805, p= 0.004), gained weight into obesity (β: 0.802, 95%: 0.409 to 1.196, p< 0.001), and those who weight cycled within/into obesity (β: 0.516, 95%: 0.021 to 1.011, p= 0.041), whilst there was little difference between the healthy weight group and those who lost weight from overweight (β: -0.039, 95%: -0.563 to 0.486, p=0.886) or from obesity (β: 0.140, 95%: -0.774 to 1.055, p= 0.764).

### Associations between weight change groups and body composition at age 62

Figure 4 and Supplementary Table S7 show the fully adjusted associations between weight change and maintenance groups at ages 50–55 and body composition at age 62. Relative to the healthy weight maintenance group, all groups - except those who lost weight from overweight - had higher FMI and FM:FFM ratio at age 62. The largest associations were observed for the obesity-related trajectory groups, particularly the obesity weight gain group (RRR: 6.673,95%: 6.143 to 7.203, p< 0.001; FM:FFM ratio: RRR: 0.275,95%: 0.245 to 0.305, p< 0.001). Although participants who lost weight from obesity also had higher FMI (RRR: 3.877,95%: 2.402 to 5.352, p= < 0.001) and FM:FFM ratio (RRR: 0.147,95%: 0.070 to 0.224, p< 0.001) than the healthy weight group, the association was smaller than the other obesity related groups, and notably smaller than for those who continued to gain weight.

**Figure 4.**
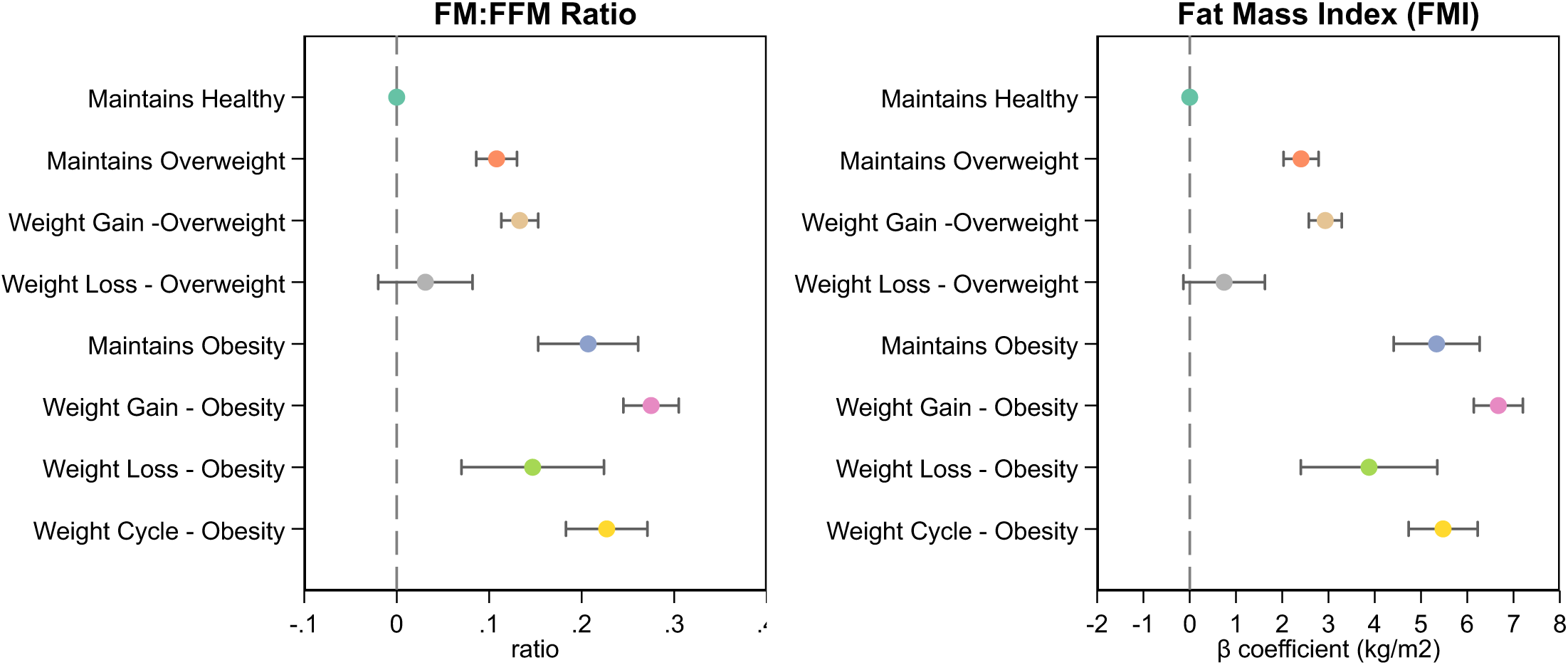
Fully adjusted Relative Risk Ratio (RRR) for association between weight change groups and body composition at age 62. *Footnote. Results from Model 2 for linear regression testing association between weight change at age 50-55 and body composition at age C2. Models are adjusted for age at outcome measurement and sex, birthweight, parental occupational social class at birth, cognitive ability at age 11, BMI and psychological distress at age 1C, adult covariates measured at age 23 (self-rated health, smoking status, occupational social class, marital status, voting behaviour, trade union membership, educational attainment, income, and membership in civic or social organisations), age peak weight was achieved up to and including age 42*.

## Discussion

Weight loss in midlife, whilst uncommon, was associated with lower cardiovascular risk across conventional cardiometabolic risk factors at age 62, while the contrasting and much more prevalent trajectory of obesity weight gain, was associated with substantially greater cardiometabolic deterioration. The benefits of weight loss were reflected in more favourable change in biomarker profiles between age 44 and 62, reflecting the change observed in the healthy weight group for triglycerides, HDLc, CRP, and systolic and diastolic blood pressure.

Individuals who lost weight in their mid-50s had both lower BMI, and improved body composition, in early old age relative to other obesity trajectories, and BMI at this later age was closely related to cardiometabolic marker levels. Together, this suggests that weight loss in later life may slow deterioration in cardiometabolic profiles through maintained reductions in BMI and improvements in body composition, bringing trajectories of biomarker change in line with those observed in individuals maintaining a healthy weight. Notably, those who continued to gain weight, particularly within or into obesity, consistently had worse biomarker profiles, higher BMI and worse body composition at age 62, and saw the fastest deterioration in cardiometabolic health markers between the ages of 44 and 62.

Whilst previous evidence indicates that an earlier onset of obesity and longer duration of exposure are associated with poorer health outcomes [3, 4], our findings suggest that meaningful improvements in cardiometabolic risk remain possible in later midlife through weight loss. Improvements in these risk factors are largely in line with corresponding changes in BMI- and adiposity-related to weight change, reflecting established links between adiposity, ectopic fat accumulation, insulin resistance, inflammation, and lipid metabolism [17–19]. In particular, we observe the most promising outcomes for those who lost weight from overweight.

Relatedly, sustained weight gain into or within overweight and obesity, as well as weight cycling within the obesity range, were associated with consistently worse biomarker trajectories relative to the healthy weight group, and broadly in line with their actual weights at 62. These findings suggest that midlife remains an important window in the life course during which to intervene [10], where preventing further weight gain could be beneficial in improve cardiometabolic and ageing outcomes. This is particularly pertinent in the context of high and increasing prevalence of obesity [6], particularly among young people [5], and the frequency in which weight gain occurs in midlife relative to much less frequent weight loss trajectories [7]. In the context of expanding availability of OMM, targeted roll out for individuals identified at high risk of continued weight gain, such as those who have experienced disadvantaged circumstances across life [7], may therefore have long-term benefits for cardiometabolic health.

Individuals who lost weight in their mid-50s exhibited a more favourable body composition at age 62, whereas those who continued to gain weight had less favourable body composition. When individuals gain fat, there are adaptive increases in lean tissue to account for the added weight [20], with concerns that weight loss may be associated with comparable mechanical reductions in fat free mass [21]. In our study, whilst FM:FFM ratios were higher among those who lost weight from obesity compared to the healthy weight group, they remained lower than the other obesity trajectories, indicating more desirable composition of fat relative to non-fat tissues. Together, these findings suggest that sustained differences in BMI and adiposity may partly explain the relationship between life-course weight trajectories and subsequent cardiometabolic risk, particularly for triglycerides, HDLc, CRP, and systolic and diastolic blood pressure.

An exception to the generally protective effects of weight loss on conventional cardiometabolic risk markers was observed for HbA1c. Elevated HbA1c levels were consistently observed among individuals with peak BMIs in the obesity range, including among those who lost weight in their 50s, when compared to individuals who had always been a healthy weight. While the association between weight loss and change in HbA1c was not statistically significant, the point estimate indicated of a potential harmful association. However, the small number of individuals who lost weight limited the precision with which this association could be estimated, resulting in large confidence intervals. Despite achieving weight loss, individuals in the obesity weight loss groups had BMIs that remained above 30kg/m^2^, indicating persistent obesity. Our study aligns with prior research in observational data that suggests HbA1c may be sensitive to cumulative duration of obesity as well as severity of obesity [3].

This study uses data from a nationally representative longitudinal study which has collected data across the life course. This meant that we were able to use repeated measures of BMI to construct weight change groups, and repeated measures of biomarkers to examine change in cardiovascular risk over time. Given the rich data available, and the known cohort properties at baseline, we were able to use multiple imputation and non-response weights to mitigate the impact of attrition and missing data [12–15].

However, interpretation of lagged associations between weight change and subsequent biomarker change as causal effects require several assumptions. Although temporally ordered in measurement, weight change between baseline and follow-up may be driven by underlying, evolving physiological or pathological processes (e.g., subclinical disease, inflammation, aging-related decline) that also continue to influence later biomarker change, creating confounding of change that is not removed by including the lagged outcome. In addition, the precise timing of weight change within the interval is unknown, so weight loss or gain may precede the first biomarker measure. Whilst adjustment for a rich set of measured confounders at age 23 can partly account for shared drivers of these changes, this assumes that such variables adequately proxy unmeasured processes. Two negative controls were used to test the adjustment set and the potential impact of residual confounding. Findings were consistent with adequate control of confounding, particularly for the weight gain groups, although the possibility of residual confounding cannot be entirely excluded. Measurement error affecting classification into weight change groups can also introduce misclassification and attenuate associations. Finally, weight change occurs within a longer life-course trajectory, and its interpretation depends on prior patterns of gain and loss. Adjustment for factors such as age at peak BMI can only partially account for this context.

Our findings have implications for two important and complementary priorities for healthy cardiometabolic ageing. First, preventing obesity, and particularly preventing continued weight gain across midlife, remains important for improving cardiometabolic risk factors, reducing accelerated cardiometabolic ageing and the growing burden on healthcare systems. Second, while prevention of obesity remains the optimal strategy, our findings suggest that there may still be cardiometabolic benefits to be gained from weight loss in later midlife. Supporting weight loss in later life could slow deterioration in multiple cardiometabolic biomarkers through reductions in achieved BMI in early old age, with little evidence of adverse effects on body composition relative to other obesity trajectories. In the context of rapidly expanding availability of OMMs, these findings suggest that supporting modest, sustained weight loss in midlife, and targeting individuals at greatest risk of continued weight gain, may represent an important opportunity to promote improved cardiometabolic ageing trajectories at the population level.

## Data Availability

The data used in this study from the National Child Development Study (NCDS) are available from the UK Data Service. Access to these data is subject to registration and may require approval from the CLS Data Access Committee to ensure the confidentiality of participants. Further information on accessing the CLS data can be found on the UK Data Service website.

https://ukdataservice.ac.uk/

## Acknowledgements

This research and the future research that these data will enable would not be possible without the valuable contributions of the NCDS cohort members over many years. We are very grateful for their ongoing commitment to the studies.

## Data Sharing Statement

The data used in this study from the National Child Development Study (NCDS) are available from the UK Data Service. Access to these data is subject to registration and may require approval from the CLS Data Access Committee to ensure the confidentiality of participants. Further information on accessing the CLS data can be found on the UK Data Service website: https://ukdataservice.ac.uk/.

## Author Contributions

CBS developed the initial idea for the paper, analysed and interpreted the data, created all figures, and drafted the manuscript. LG derived variables for analysis, and edited the manuscript, and all authors (LG, NS, NC, GBP) critically revised the manuscript for intellectual content and gave final approval for the version published.

## References

1. Aune, D., et al., BMI and all cause mortality: systematic review and non-linear dose-response meta-analysis of 230 cohort studies with 3.74 million deaths among 30.3 million participants. British Medical Journal, 2016. 353: i2156.

2. Abdelaal, M., C.W. le Roux, and N.G. Docherty, Morbidity and mortality associated with obesity. Annals of Translational Medicine, 2017. 5(7): 161.

3. Norris, T., et al., Duration of obesity exposure between ages 10 and 40 years and its relationship with cardiometabolic disease risk factors: A cohort study. PLOS Medicine, 2020. 17(12): e1003387.

4. Le, H.T., et al., Weight trajectories and obesity onset between 17 and 60 years of age, and cause-specific mortality: the Obesity and Disease Development Sweden (ODDS) pooled cohort study. EClinicalMedicine, 2026. 94: 103870.

5. Fletcher, R.A., et al., Whole-population trends in obesity across dimensions of inequality in England, 2019-25: a retrospective, longitudinal cohort study of 54 million adults. Lancet Diabetes & Endocrinology, 2026. 14: 740–753

6. Johnson, W., et al., How Has the Age-Related Process of Overweight or Obesity Development Changed over Time? Co-ordinated Analyses of Individual Participant Data from Five United Kingdom Birth Cohorts. PLOS Medicine, 2015. 12(5): e1001828.

7. Bridger Staatz, C., L. Gimeno, D. Smeeth, N. Sattar, N. Chaturvedi, and G.B. Ploubidis, Hard to lose, easy to gain; Trends in obesity and weight change across the life course in four British Birth cohorts. [Preprint] medRxiv 2026. 10.64898/2026.06.22.26356252

8. Gimeno, L., et al., The generational health drift: A systematic review of evidence from the British birth cohort studies. Population Studies, 2026. 1–19.

9. Jackson, S.E., et al., Prevalence of use and interest in using glucagon-like peptide-1 receptor agonists for weight loss: a population study in Great Britain. BMC Medicine, 2026. 24(1): 1.

10. Lachman, M.E., S. Teshale, and S. Agrigoroaei, Midlife as a pivotal period in the life course: Balancing growth and decline at the crossroads of youth and old age. International Journal of Behavioral Development, 2015. 39(1): 20–31.

11. Power, C. and J. Elliott, Cohort profile: 1958 British Birth Cohort (National Child Development Study). International Journal of Epidemiology, 2006. 35(1): 34–41.

12. Mostafa, T., et al., Missing at random assumption made more plausible: evidence from the 1958 British birth cohort. Journal of Clinical Epidemiology, 2021. 136: p. 44–54.

13. Silverwood, R., Narayanan, M., Dodgeon, B., Katsoulis, M., Ploubidis, G, Handling missing data in the CLS cohort studies: User guide. 2024: London: UCL Centre for Longitudinal Studies.

14. Narayanan, M.K., et al., How to mitigate selection bias in COVID-19 surveys: evidence from five national cohorts. European Journal of Epidemiology, 2024. 39(11): 1221–1227.

15. Katsoulis, M., et al., A data driven approach to address missing data in the 1970 British birth cohort. BMC Medical Research Methodology, 2026. 26(1): 91.

16. Lipsitch, M., E.T. Tchetgen, and T. Cohen, Negative Controls: A Tool for Detecting Confounding and Bias in Observational Studies. Epidemiology, 2010. 21(3): 383–388.

17. Powell-Wiley, T.M., et al., Obesity and Cardiovascular Disease: A Scientific Statement From the American Heart Association. Circulation, 2021. 143(21): e984–e1010.

18. Neeland, I.J., et al., Visceral and ectopic fat, atherosclerosis, and cardiometabolic disease: a position statement. Lancet Diabetes & Endocrinology, 2019. 7(9): 715–725.

19. Sattar, N. and J.M.R. Gill, Type 2 diabetes as a disease of ectopic fat? BMC Medicine, 2014. 12: 123.

20. Forbes, G.B., Some adventures in body composition, with special reference to nutrition. Acta Diabetologica, 2003. 40: S238–S241.

21. Caturano, A., et al., Sarcopenic obesity and weight loss-induced muscle mass loss. Current Opinion in Clinical Nutrition & Metabolic Care, 2025. 28(4): 339–350.

