## Supplementary Material for "Associations between adult obesity and mid-life weight change patterns with cardiometabolic health in early old age: Evidence from the 1958 British birth cohort"

### Supplementary Materials

#### S1. Derivation of biomarkers

| **Biomarker** | **Derivation** |
|---|---|
| Systolic and diastolic blood pressure | At age 44, mean systolic and diastolic blood pressure measured using Omron 907 blood pressure monitor, across two readings, while seated. At age 62, measured using Omron HEM 907 blood pressure monitor, across two readings, while seated.<br><br>Systolic blood pressure < 70 mmHg or > 270 mmHg were considered implausible and set to missing (0 cases at age 44, 0 cases at age 62), as were diastolic blood pressure measures < 40 mmHg (1 case at age 44, 1 case at age 62). [1]<br><br>For those taking medication for high blood pressure, including diuretics, beta-blockers and medication for heart failure (BNF chapters 2.02, 2.04 and 2.05), we added a +10 mmHg correction to systolic blood pressure, and a +5 mmHg correction to diastolic blood pressure. [2, 3] |
| Total cholesterol | At both ages 44 and 62, from non-fasted serum, from blood samples excluding people with clotting/bleeding disorders, on anticoagulant therapy, who had had a fit in the last 3 years, or who were not willing to give written consent. At age 44, measured using Olympus AU640 autoanalyser. At age 62, measured using Roche Cobas c702 generation 4 assay.<br><br>Values are given in mmol/L. Values <1.75 or >20 mmol/L were considered implausible and set to missing (0 cases at age 44; 0 cases at age 62). [4]<br><br>For those taking lipid-regulating drugs, including statins (BNF chapter 2.12), we multiplied observed total cholesterol values by 1.25. [5] |
| HDL cholesterol | At both ages 44 and 62, from non-fasted serum, from blood samples excluding people with clotting/bleeding disorders, on anticoagulant therapy, who had had a fit in the last 3 years, or who were not willing to give written consent. At age 44, measured using Olympus AU640 auto-analyser. At age 62, measured using Roche Cobas c702 generation 4 assay.<br><br>Values are given in mmol/L. Values <0.4 or >5 mmol/L were considered implausible and set to missing (0 cases at age 44; 0 cases at age 62). [4]<br><br>For those taking lipid-regulating drugs, including statins (BNF chapter 2.12), we multiplied observed HDL cholesterol values by 0.75. [5]<br><br>See Text S8 for a discussion of HDLc as a marker of CVD risk. |

| | |
|---|---|
| LDL cholesterol | While total cholesterol, HDL cholesterol and triglyceride levels were measured directly, we derived LDL cholesterol levels using the Martin Hopkins formula. [6]<br><br>For those taking lipid-regulating drugs, including statins (BNF chapter 2.12), we multiplied observed LDL cholesterol values by 1.54. [5] |
| Triglycerides | At both ages 44 and 62, from non-fasted serum, from blood samples excluding people with clotting/bleeding disorders, on anticoagulant therapy, who had had a fit in the last 3 years, or who were not willing to give written consent. At age 44, measured using Olympus AU640 auto-analyser. At age 62, measured using Roche Cobas c702.<br><br>Triglyceride values <0.11 mmol/L or >33.3 mmol/L were considered implausible and set to missing (0 cases at age 44; 0 cases at age 62). [7]<br><br>For those taking lipid-regulating drugs, including statins (BNF chapter 2.12), we multiplied measured triglyceride levels by 1.18. [8, 5] |
| HbA1c | At age 44, measured on whole citrated blood by ion exchange high-performance liquid chromatography (HPLC) using Tosoh A1c 2.2 Glycohemoglobin Analyser HLC-723GHb. At age 62, also measured using ion exchange HPLC using Tosoh G8/G11.<br><br>Values are given in DCCT (%) units. Values > 20.4% were considered implausible and set to missing (0 cases at age 44, 0 cases at age 62). [9]<br><br>For those taking medication for diabetes (BNF chapter 6.1), we added a +1 percentage-point correction to the measured HbA1c value. [9] |
| CRP | At age 44, measured on citrated plasma by high sensitivity nephelometric analysis of latex particles coated with CRP monoclonal antibodies. At age 62, measured using particle-enhanced immunoturbidimetry using Roche Cobas c702.<br><br>Values > 10 mg/L were set as missing, as these are likely to indicate infection (225 cases at age 44, 165 cases at age 62). [10] |
| FMI | Body fat percentage (BF%) was measured by Tanita BF - 522W scales using foot-to-foot bioelectrical impedance analysis at age 62. The scales can accurately measure weight up to 130kg, and those who would likely exceed this weight were not weighed. |

| | |
|---|---|
| | Respondents' age, gender and height were entered into the scales before measuring, to ensure accuracy. BF % is calculated as total fat weight as a percentage of total body weight. Using BF% it is possible to derive total fat mass, and then calculated fat mass index (FMI) as FMI = fat mass (kg) / height (m) $^2$ |
| FM:FFM Ratio | Once fat mass (FM) has been derived, fat-free mass (FFM) can be derived: FFM (kg) = body weight (kg) - FM (kg).<br>The ratio of FM and FFM can then be calculated: FM:FFM = FM/FFM. |
| Heart Rate (Pulse) | At age 44, pulse (beats per minute) was measured using Omron 907 blood pressure monitor, across three readings, while seated. At age 62, pulse (beats per minute) was measured using Omron HEM 907 blood pressure monitor, across two readings, while seated.<br><br>The mean pulse per minute were calculated at age 44 and 62, from the three and two readings, respectively. |

[1] Devereux RB, James GD, Pickering TG. What is normal blood pressure? Comparison of ambulatory pressure level and variability in patients with normal and abnormal left ventricular geometry. *Am J Hypertension,* 1993; 6:211S-215S. https://doi.org/10.1093/ajh/6.6.211S

[2] Law MR, Wald NJ, Morris JK, Jordan RE. Value of low dose combination treatment with blood pressure lowering drugs: Analysis of 354 randomised trials. *BMJ,* 2003; 326:1427. https://doi.org/10.1136/bmj.7494.1427

[3] Tobin MD, Sheehan NA, Scurrah KJ, Burton PR. Adjusting for treatment effects in studies of quantitative traits: antihypertensive therapy and systolic blood pressure. *Stat Med,* 2005; 24(19): 2911-2935. https://doi.org/10.1002/sim.2165

[4] NCD Risk Factor Collaboration (NCD-RisC). National trends in total cholesterol obscure heterogeneous changes in HDL and non-HDL cholesterol and total-to-HDL cholesterol ratio: A pooled analysis of 458 population-based studies and Asian and Western countries. *Int J Epidemiol,* 2020; 49(1):173-192. https://doi.org/10.1093/ije/dyz099

[5] Ploubidis GB, Batty GD, Patalay P, Bann D, Goodman A. Association of early-life mental health with biomarkers in midlife and premature mortality: Evidence from the 1958 British birth cohort. *JAMA Psychiatry,* 2021; 78(1): 38-46. https://doi.org/10.1001/jamapsychiatry.2020.2893

[6] Martins J, Steyn N, Rossouw HM*, et al.* Best practice for LDL-cholesterol: when and how to calculate. *J Clin Pathol* 2023; **76:** 145-152. https://doi.org/10.1136/jcp-2022-208480

[7] Cheng FW, Gao X, Bao L *et al.* Obesity as a risk factor for developing functional limitation among older adults. A conditional inference tree analysis. *Obesity*, 2017; 25(7): 1147-1292. https://doi.org/10.1002/oby.21861

[8] Batty GD, Hamer M. Public care during childhood and biomedical risk factors in middle age: The 1970 British Cohort Study. *Am J Epidemiol,* 2021; 190(1):176-178. https://doi.org/10.1093/aje/kwaa079

[9] Garfield B, Farmaki AE, Eastwood SV, *et al*. HbA1c and brain health across the entire glycaemic spectrum. *Diabetes Obesity Metabolism,* 2022; 24(7): 1406. https://doi.org/10.1111/dom.14321

[10] Ruiz M, Benzeval M, Kumari M. *A guide to the biomarker data in the CLOSER studies*. 2017. London: Cohort and Longitudinal Studies Enhancement Recourses. pp. 104.

#### S2. Derivation of weight change and maintenance groups

We defined weight loss in the final two sweeps of NCDS (age 50 and 55), known as the 'observation period', as weight loss of greater than 5% compared to the highest recorded weight in adulthood (peak weight) observed between the ages of 23 and 42.

**Maintained normal weight.** Individuals who had a healthy (or underweight) peak BMI (≤25 k/m2) between ages 23 and 42 and maintained a healthy (or underweight) BMI during the observation period. Some individuals in this group may lose or gain weight across the life course, but their BMI remains ≤25kg/$m^2$ at each observation.

**Mostly maintained overweight.** Individuals who had an overweight peak BMI (>25kg/$m^2$ and ≤30kg/$m^2$) between ages 23 and 42 and/or and maintained this weight without additional weight gain (>5% of body weight) or weight loss (>5% of body weight). This group also included individuals who lost more than 5% of body weight in the final observation (at age 55 ) only, but for whom 'sustained' weight loss could not be confirmed.

This group also included individuals whose peak BMI was healthy but moved into the overweight group without substantial weight gain (>5%) in either of the final two observations, or the alternative where their weight reduced by less than 5% from peak, which may have moved them to a healthy weight. Individuals who weight cycled (see fluctuations in their weight) but experience one observation in the overweight category at either at peak BMI or in the observation window are also included in this group.

**Weight gain into overweight.** This group includes people who gained more than 5% of their body weight in the final observation, relative to their peak BMI before age 42, and their final BMI is consider overweight (>25kg/$m^2$ and ≤30kg/$m^2$)

**Weight loss from overweight.** This group includes people who 'sustained >5% weight loss', but from a peak BMI >25kg/$m^2$ and ≤30kg/$m^2$ at both time points in the observation window. This group includes both individuals whose weight loss returned their weight to healthy in the final two observations or remained in the overweight category.

**Mostly maintained obesity.** Individuals who had an obese peak BMI (>30kg/$m^2$) between ages 23 and 42 and maintained this weight without additional weight gain (>5% of body weight) or weight loss (>5% of body weight). This group also included individuals who lost more than 5% of body weight in the final observation (at age 55) only, but for whom 'sustained' weight loss could not be confirmed.

This group also contained several marginal cases, where no significant weight loss or gain occurred (>5%) between peak BMI and the observational period, but peak BMI or final BMI was obese (>30kg/$m^2$).

**Obese – gain >5% of body weight.** Individuals who gained more than 5% of body weight compared to their previously highest recorded weight, and whose final BMI was >30kg/$m^2$. This category could also include individuals that were previously healthy weight or overweight based on their peak BMI, but who gained weight to end up with an obese BMI at age 55 , as well as individuals who were already overweight or obese, and further increased their weight >5%.

**Obese – weight loss >5% of body weight.** Individuals who lost more than 5% of body weight at both observations in the final two sweeps compared to a previous obese peak BMI (>30kg/$m^2$) between ages 23 and 42. Weight loss may occur within the obesity BMI class or take cohort members from an obese BMI to an overweight or healthy BMI. This group excludes individuals who lost >5% of body weight at only one of the two final observations.

**Obese – weight cycling.** Broadly speaking, these were individuals who either lost or gained 5% relative to their peak weight between ages 23and 42, but for who this was only observed at one of the two final observations (i.e., not sustained). For these individuals, their BMI was >30kg/$m^2$ at either peak, age 50 or 55, or at multiple observations.

More specifically, this group included:

- Individuals that experienced 5% weight loss at age 50 compared to their peak weight but returned to within 5% of peak weight at the final observation, and who had an obese final BMI.
- Individuals that experienced 5% weight loss at age 50 compared to their peak weight, then gained an additional 5% at age 55 compared to their peak weight, and who were obese at either peak or the final observation.
- Individuals that experienced 5% weight gain at age 50, but whose BMI returned to within 5% of peak by the final observation and had an obese BMI during the observation period.

- Individuals that experienced 5% weight gain at age 50, then experienced 5% weight loss in the final observation compared to their peak BMI and had an obese BMI at either peak or the final observation.
- Individuals whose weight was 5% higher compared to peak at both ages 50 and 55. Notable weight gain occurred at age 50, and weight was lost relative to age 50 but not peak BMI. These individuals experienced a large amount of weight gain at age 50, taking them from normal/borderline overweight (peak BMI) to obese (age 50) to overweight (age 55).
- Individuals whose weight was 5% lower than peak at age 50, having come from marginally obese peak BMI but then returning to within 5% of peak by age 55.

Given the intervals between sweeps it is likely that there are “weight cyclers” in the other groups, but we were unable to identify them with the available data.

We did not use nurse-measured BMI at age 44 in NCDS to define lifetime prevalence groups, peak BMI, or subsequent weight loss, to avoid measurement mode effect. Including nurse-measured BMI as part of the observations used to define peak BMI would risk overestimating weight loss, since subsequent BMI observations used to identify weight loss were based on self-reports, which likely underestimate true BMI.

#### S3. Auxiliary variables

Multiple imputation works under the assumption that data are Missing At Random (MAR), that is, that missingness is random conditional on observed characteristics. Auxiliary variables can be added to multiple imputation models to increase the plausibility of this untestable assumption. Auxiliary variables included in the imputation model are shown below, and were informed by previous research on predictors of sweep response in NCDS:

*Early life characteristics*

- Country of birth (age 0)
- Birthweight (age 0)
- Number of births mother had already had (age 0)
- Whether mother stayed to minimum school leaving age (age 0)
- Whether mother smoked during pregnancy (age 0)
- Number of rooms in accommodation (age 0)
- Residential movement in childhood (age 7)
- Housing tenure (age 11)
- Cognitive ability (age 11)
- Medical conditions in childhood (age 11)
- Breastfeeding in early life (reported by mother when CM aged 7)
- Whether parents read to the CM in childhood (age 7)
- Psychological distress in adolescence (age 16)

*Adult characteristics*

- Psychological distress (age 33)
- Presence of longstanding illness (age 33 and 42)
- Smoking status (age 23 and 42)
- Self-rated health (age 23 and 42)
- Life satisfaction (age 42)
- Housing tenure (age 33)
- Household income (age 23)
- Manual occupational social class (age 42)
- Partnership status (age 23, 33 and 42)
- Social trust (age 62)
- Voting (age 23 and 42)

- Union membership (age 23, 33 and 42)
- Membership in civic or social organisations (age 23)
- Whether lived alone (age 42)
- Access to social support (age 42)
- Cognition (age 50)
- Additional biomarkers at age 44: Growth differentiation factor-15 (GDF-15), N-terminal prohormone (NT-proBNP), and High sensitivity Troponin-I (hs-cTnI)
- An indicator of response to age 44 biomedical sweep
- Number of sweeps completed (age 62)

#### S4. Mean BMI by weight change and maintenance groups

**Supplementary Table S4. Mean BMI from age 16 to age 62 by weight change groups defined at ages 50 to 55.**

| Mean (SE) BMI by age and weight change group | Age 16 | Age 23 | Age 33 | Age 42 | Age 50 | Age 55 | Age 62 |
|---|---|---|---|---|---|---|---|
| Maintains healthy (ref) | 19.26 (0.11) | 20.24 (0.13) | 21.06 (0.11) | 21.48 (0.10) | 22.02 (0.11) | 22.15 (0.11) | 23.08 (0.14) |
| Maintains overweight | 20.61 (0.16) | 22.67 (0.14) | 24.94 (0.16) | 25.94 (0.13) | 26.79 (0.11) | 26.01 (0.12) | 27.31 (0.18) |
| Weight Gain – Overweight | 19.80 (0.15) | 21.45 (0.13) | 23.19 (0.15) | 24.08 (0.12) | 26.11 (0.12) | 27.54 (0.07) | 27.80 (0.18) |
| Weigh Loss – Overweight | 21.20 (0.37) | 23.15 (0.31) | 25.84 (0.38) | 25.69 (0.33) | 24.26 (0.29) | 23.84 (0.30) | 25.05 (0.49) |
| Maintains Obesity | 22.75 (0.27) | 26.06 (0.30) | 30.81 (0.37) | 32.64 (0.35) | 33.28 (0.33) | 32.22 (0.37) | 33.20 (0.51) |
| Weight Gain – Obesity | 21.44 (0.17) | 23.95 (0.17) | 27.49 (0.23) | 29.27 (0.24) | 32.72 (0.25) | 34.97 (0.24) | 34.04 (0.26) |
| Weight Loss – Obesity | 23.57 (0.71) | 26.78 (0.66) | 32.62 (0.92) | 32.51 (0.85) | 30.14 (0.64) | 30.05 (0.75) | 31.07 (0.89) |
| Weight Cycle - Obesity | 22.36 (0.30) | 25.13 (0.32) | 29.04 (0.40) | 30.83 (0.40) | 33.18 (0.46) | 31.61 (0.37) | 32.70 (0.45) |

#### S5. Associations between weight change and maintenance groups at 50-55 and lagged biomarkers at 62

| | **Model 1 – sex and age adjusted** | | **Model 2 – M1 + lagged outcome** | | **Model 3 – M2 + covariates + peak BMI age (+ all biomarkers)** | |
|---|---|---|---|---|---|---|
| | **Coefficient (95% CI)** | **P Value** | **Coefficient (95% CI)** | **P Value** | **Coefficient (95% CI)** | **P Value** |
| ***Systolic BP*** | | | | | | |
| Maintains healthy (ref) | - | - | - | - | - | - |
| Maintains overweight | 4.699 (1.887 to 7.511) | 0.001 | 2.577 (-0.194 to 5.347) | 0.068 | 3.348 (0.477 to 6.219) | 0.022 |
| Weight Gain – Overweight | 4.818 (1.524 to 8.112) | 0.004 | 3.109 (0.146 to 6.072) | 0.04 | 3.522 (0.625 to 6.418) | 0.017 |
| Weigh Loss – Overweight | 3.823 (-3.686 to 11.332) | 0.318 | 1.374 (-5.400 to 8.147) | 0.691 | 2.234 (-4.655 to 9.124) | 0.525 |
| Maintains Obesity | 7.037 (2.990 to 11.084) | 0.001 | 1.193 (-2.808 to 5.195) | 0.559 | 3.028 (-1.501 to 7.558) | 0.19 |
| Weight Gain – Obesity | 8.141 (5.272 to 11.011) | < 0.001 | 4.334 (1.721 to 6.947) | 0.001 | 5.726 (2.660 to 8.793) | < 0.001 |
| Weight Loss – Obesity | 3.458 (-4.151 to 11.068) | 0.373 | -1.336 (-8.404 to 5.732) | 0.711 | 0.947 (-6.605 to 8.499) | 0.806 |
| Weight Cycle - Obesity | 7.099 (2.812 to 11.387) | 0.001 | 2.318 (-1.628 to 6.264) | 0.249 | 4.092 (-0.318 to 8.502) | 0.069 |
| ***Diastolic* BP** | | | | | | |
| Maintains healthy (ref) | - | - | - | - | - | - |
| Maintains overweight | 2.788 (1.366 to 4.209) | < 0.001 | 1.541 (0.152 to 2.931) | 0.03 | 2.050 (0.521 to 3.580) | 0.009 |
| Weight Gain – Overweight | 2.379 (0.696 to 4.062) | 0.006 | 1.420 (-0.175 to 3.016) | 0.081 | 1.812 (0.224 to 3.401) | 0.025 |
| Weigh Loss – Overweight | 1.640 (-2.589 to 5.870) | 0.447 | 0.396 (-3.533 to 4.325) | 0.844 | 1.109 (-2.908 to 5.126) | 0.588 |
| Maintains Obesity | 3.677 (1.039 to 6.315) | 0.006 | 0.449 (-2.243 to 3.141) | 0.744 | 2.064 (-0.812 to 4.940) | 0.16 |
| Weight Gain – Obesity | 4.564 (2.895 to 6.234) | < 0.001 | 2.336 (0.725 to 3.947) | 0.004 | 3.519 (1.703 to 5.335) | < 0.001 |
| Weight Loss – Obesity | 1.379 (-2.956 to 5.714) | 0.533 | -0.753 (-4.656 to 3.149) | 0.705 | 1.367 (-2.823 to 5.556) | 0.522 |
| Weight Cycle - Obesity | 3.595 (1.054 to 6.137) | 0.006 | 1.058 (-1.449 to 3.564) | 0.408 | 2.461 (-0.305 to 5.228) | 0.081 |
| ***HDL Cholesterol*** | | | | | | |
| Maintains healthy (ref) | - | - | - | - | - | - |
| Maintains overweight | -0.213 (-0.284 to -0.141) | < 0.001 | -0.114 (-0.174 to -0.055) | < 0.001 | -0.084 (-0.143 to -0.025) | 0.005 |
| Weight Gain – Overweight | -0.255 (-0.332 to -0.177) | < 0.001 | -0.159 (-0.225 to -0.093) | < 0.001 | -0.126 (-0.188 to -0.064) | < 0.001 |
| Weigh Loss – Overweight | -0.097 (-0.236 to 0.041) | 0.167 | -0.028 (-0.145 to 0.089) | 0.637 | -0.004 (-0.117 to 0.109) | 0.946 |
| Maintains Obesity | -0.424 (-0.533 to -0.315) | < 0.001 | -0.214 (-0.302 to -0.125) | < 0.001 | -0.145 (-0.242 to -0.047) | 0.004 |
| Weight Gain – Obesity | -0.451 (-0.524 to -0.377) | < 0.001 | -0.252 (-0.321 to -0.182) | < 0.001 | -0.189 (-0.259 to -0.118) | < 0.001 |
| Weight Loss – Obesity | -0.326 (-0.579 to -0.072) | 0.012 | -0.137 (-0.342 to 0.068) | 0.191 | -0.072 (-0.273 to 0.128) | 0.479 |
| Weight Cycle - Obesity | -0.416 (-0.525 to -0.308) | < 0.001 | -0.221 (-0.318 to -0.125) | < 0.001 | -0.152 (-0.254 to -0.050) | 0.003 |
| ***LDL Cholesterol*** | | | | | | |
| Maintains healthy (ref) | - | - | - | - | - | - |
| Maintains overweight | 0.152 (-0.005 to 0.308) | 0.058 | 0.027 (-0.116 to 0.170) | 0.71 | 0.060 (-0.081 to 0.200) | 0.403 |

| | | | | | | |
|---|---|---|---|---|---|---|
| Weight Gain – Overweight | 0.234 (0.046 to 0.422) | 0.015 | 0.105 (-0.062 to 0.271) | 0.218 | 0.120 (-0.050 to 0.290) | 0.166 |
| Weigh Loss – Overweight | -0.044 (-0.393 to 0.304) | 0.803 | -0.102 (-0.424 to 0.221) | 0.536 | -0.050 (-0.366 to 0.266) | 0.756 |
| Maintains Obesity | -0.116 (-0.397 to 0.165) | 0.42 | -0.294 (-0.566 to -0.021) | 0.034 | -0.205 (-0.480 to 0.069) | 0.142 |
| Weight Gain – Obesity | 0.086 (-0.095 to 0.267) | 0.353 | -0.114 (-0.285 to 0.056) | 0.188 | -0.055 (-0.246 to 0.136) | 0.57 |
| Weight Loss – Obesity | -0.048 (-0.506 to 0.409) | 0.836 | -0.194 (-0.620 to 0.232) | 0.372 | -0.078 (-0.511 to 0.355) | 0.724 |
| Weight Cycle - Obesity | 0.045 (-0.183 to 0.273) | 0.701 | -0.169 (-0.378 to 0.039) | 0.112 | -0.093 (-0.323 to 0.138) | 0.431 |
| ***Heart Rate (Pulse)*** | | | | | | |
| Maintains healthy (ref) | - | - | - | - | - | - |
| Maintains overweight | -0.087 (-1.681 to 1.507) | 0.915 | -0.400 (-1.814 to 1.014) | 0.579 | -0.196 (-1.587 to 1.194) | 0.782 |
| Weight Gain – Overweight | -0.123 (-1.712 to 1.467) | 0.88 | -0.503 (-1.999 to 0.993) | 0.51 | -0.359 (-1.863 to 1.145) | 0.64 |
| Weigh Loss – Overweight | -0.746 (-4.285 to 2.793) | 0.68 | -0.884 (-3.931 to 2.164) | 0.57 | -0.983 (-3.988 to 2.022) | 0.521 |
| Maintains Obesity | 2.622 (-0.498 to 5.743) | 0.1 | 0.970 (-1.945 to 3.886) | 0.514 | 1.191 (-2.011 to 4.393) | 0.466 |
| Weight Gain – Obesity | 1.876 (0.116 to 3.635) | 0.037 | 0.369 (-1.255 to 1.993) | 0.656 | 0.576 (-1.292 to 2.443) | 0.546 |
| Weight Loss – Obesity | 1.537 (-3.262 to 6.335) | 0.53 | 0.372 (-3.789 to 4.534) | 0.861 | 0.702 (-3.740 to 5.145) | 0.757 |
| Weight Cycle - Obesity | 1.302 (-0.926 to 3.529) | 0.252 | -0.017 (-2.163 to 2.128) | 0.987 | 0.297 (-2.014 to 2.609) | 0.801 |
| ***Triglycerides*** | | | | | | |
| Maintains healthy (ref) | - | - | - | - | - | - |
| Maintains overweight | 0.228 (0.076 to 0.381) | 0.003 | 0.111 (-0.047 to 0.269) | 0.17 | 0.087 (-0.075 to 0.249) | 0.294 |
| Weight Gain – Overweight | 0.389 (0.194 to 0.584) | < 0.001 | 0.282 (0.080 to 0.484) | 0.006 | 0.229 (0.047 to 0.411) | 0.013 |
| Weigh Loss – Overweight | 0.040 (-0.290 to 0.370) | 0.812 | -0.057 (-0.399 to 0.284) | 0.743 | -0.051 (-0.392 to 0.290) | 0.77 |
| Maintains Obesity | 0.478 (0.190 to 0.766) | 0.001 | 0.127 (-0.160 to 0.414) | 0.386 | 0.111 (-0.185 to 0.406) | 0.463 |
| Weight Gain – Obesity | 0.706 (0.478 to 0.934) | < 0.001 | 0.432 (0.199 to 0.664) | < 0.001 | 0.366 (0.118 to 0.614) | 0.004 |
| Weight Loss – Obesity | 0.416 (-0.154 to 0.985) | 0.153 | 0.161 (-0.375 to 0.697) | 0.557 | 0.145 (-0.361 to 0.650) | 0.575 |
| Weight Cycle - Obesity | 0.506 (0.252 to 0.761) | < 0.001 | 0.215 (-0.081 to 0.510) | 0.155 | 0.171 (-0.131 to 0.474) | 0.267 |
| ***HbA1c*** | | | | | | |
| Maintains healthy (ref) | - | - | - | - | - | - |
| Maintains overweight | 0.214 (0.061 to 0.367) | 0.006 | 0.132 (0.001 to 0.263) | 0.047 | 0.065 (-0.067 to 0.197) | 0.333 |
| Weight Gain – Overweight | 0.195 (0.036 to 0.354) | 0.016 | 0.160 (0.026 to 0.294) | 0.019 | 0.078 (-0.055 to 0.211) | 0.253 |
| Weigh Loss – Overweight | 0.210 (-0.069 to 0.489) | 0.14 | 0.046 (-0.196 to 0.288) | 0.709 | 0.019 (-0.223 to 0.262) | 0.876 |
| Maintains Obesity | 1.026 (0.624 to 1.428) | < 0.001 | 0.655 (0.305 to 1.005) | < 0.001 | 0.470 (0.096 to 0.845) | 0.014 |
| Weight Gain – Obesity | 0.756 (0.563 to 0.949) | < 0.001 | 0.553 (0.386 to 0.721) | < 0.001 | 0.372 (0.179 to 0.565) | < 0.001 |
| Weight Loss – Obesity | 1.255 (0.412 to 2.097) | 0.004 | 0.582 (-0.080 to 1.245) | 0.085 | 0.475 (-0.187 to 1.137) | 0.16 |
| Weight Cycle - Obesity | 0.757 (0.460 to 1.053) | < 0.001 | 0.474 (0.225 to 0.723) | < 0.001 | 0.306 (0.031 to 0.580) | 0.029 |
| ***C-Reactive Protein*** | | | | | | |
| Maintains healthy (ref) | - | - | - | - | - | - |

| | | | | | | |
|---|---|---|---|---|---|---|
| Maintains overweight | 0.424 (0.114 to 0.734) | 0.007 | 0.234 (-0.057 to 0.525) | 0.115 | 0.206 (-0.094 to 0.506) | 0.178 |
| Weight Gain – Overweight | 0.697 (0.376 to 1.017) | < 0.001 | 0.528 (0.217 to 0.838) | 0.001 | 0.478 (0.152 to 0.805) | 0.004 |
| Weigh Loss – Overweight | 0.137 (-0.450 to 0.724) | 0.648 | 0.001 (-0.568 to 0.570) | 0.998 | -0.039 (-0.563 to 0.486) | 0.886 |
| Maintains Obesity | 1.042 (0.381 to 1.702) | 0.002 | 0.452 (-0.190 to 1.094) | 0.167 | 0.410 (-0.306 to 1.127) | 0.261 |
| Weight Gain – Obesity | 1.433 (1.052 to 1.813) | < 0.001 | 0.882 (0.500 to 1.264) | < 0.001 | 0.802 (0.409 to 1.196) | < 0.001 |
| Weight Loss – Obesity | 0.792 (-0.119 to 1.702) | 0.088 | 0.145 (-0.721 to 1.011) | 0.743 | 0.140 (-0.774 to 1.055) | 0.764 |
| Weight Cycle - Obesity | 1.169 (0.663 to 1.676) | < 0.001 | 0.588 (0.078 to 1.098) | 0.024 | 0.516 (0.021 to 1.011) | 0.041 |

**Table S5 Footnotes:** Full model results for lagged linear regression models exploring associations between weight change groups at age 50-55 and biomarkers at age 62, showing rate of change in biomarker between age 44 and 62. Model 1 adjusted for age at outcome measurement and sex only. Model 2 additionally adjusted for the corresponding biomarker measured at age 44. Model 3 additionally adjusted for birthweight, parental occupational social class at birth, cognitive ability at age 11, BMI and psychological distress at age 16, adult covariates measured at age 23 (self-rated health, smoking status, occupational social class, marital status, voting behaviour, trade union membership, educational attainment, income, and membership in civic or social organisations), age peak weight was achieved up to and including age 42, and all other biomarker measures at age 44.

### S6. Associations between weight change and maintenance groups at 50-55 and body composition at age 62

| | **Model 1 – sex and age adjusted** | | **Model 2 – Model 1 + covariates + peak BMI age** | |
|---|---|---|---|---|
| | **Coefficient (95% CI)** | **P Value** | **Coefficient (95% CI)** | **P Value** |
| ***FMI*** | | | | |
| Maintains healthy (ref) | - | - | - | - |
| Maintains overweight | 2.803 (2.418 to 3.188) | < 0.001 | 2.406 (2.027 to 2.785) | < 0.001 |
| Weight Gain – Overweight | 3.212 (2.871 to 3.553) | < 0.001 | 2.930 (2.574 to 3.286) | < 0.001 |
| Weigh Loss – Overweight | 1.126 (0.231 to 2.022) | 0.014 | 0.744 (-0.138 to 1.627) | 0.098 |
| Maintains Obesity | 6.240 (5.291 to 7.190) | < 0.001 | 5.338 (4.409 to 6.267) | < 0.001 |
| Weight Gain – Obesity | 7.340 (6.819 to 7.862) | < 0.001 | 6.673 (6.143 to 7.203) | < 0.001 |
| Weight Loss – Obesity | 4.760 (3.270 to 6.249) | < 0.001 | 3.877 (2.402 to 5.352) | < 0.001 |
| Weight Cycle - Obesity | 6.285 (5.535 to 7.035) | < 0.001 | 5.478 (4.732 to 6.225) | < 0.001 |
| ***FM:FFM Ratio*** | | | | |
| Maintains healthy (ref) | - | - | - | - |
| Maintains overweight | 0.124 (0.101 to 0.146) | < 0.001 | 0.108 (0.086 to 0.130) | < 0.001 |
| Weight Gain – Overweight | 0.146 (0.126 to 0.166) | < 0.001 | 0.133 (0.113 to 0.153) | < 0.001 |
| Weigh Loss – Overweight | 0.045 (-0.006 to 0.095) | 0.083 | 0.031 (-0.020 to 0.082) | 0.232 |
| Maintains Obesity | 0.242 (0.188 to 0.296) | < 0.001 | 0.207 (0.153 to 0.261) | < 0.001 |
| Weight Gain – Obesity | 0.304 (0.274 to 0.334) | < 0.001 | 0.275 (0.245 to 0.305) | < 0.001 |
| Weight Loss – Obesity | 0.180 (0.105 to 0.255) | < 0.001 | 0.147 (0.070 to 0.224) | < 0.001 |
| Weight Cycle - Obesity | 0.259 (0.215 to 0.303) | < 0.001 | 0.227 (0.183 to 0.271) | < 0.001 |

**Table S7 Footnotes:** Full model results for linear regression models exploring associations between weight change groups at age 50-55 and body composition at age 62, without a corresponding lagged variable available at age 44. Model 1 adjusted for age at outcome measurement and sex only. Model 2 additionally adjusted for for birthweight, parental occupational social class at birth, cognitive ability at age 11, BMI and psychological distress at age 16, adult covariates measured at age 23 (self-rated health, smoking status, occupational social class, marital status, voting behaviour, trade union membership, educational attainment, income, and membership in civic or social organisations) and age peak weight was achieved up to and including age 42.

#### S7. Negative controls

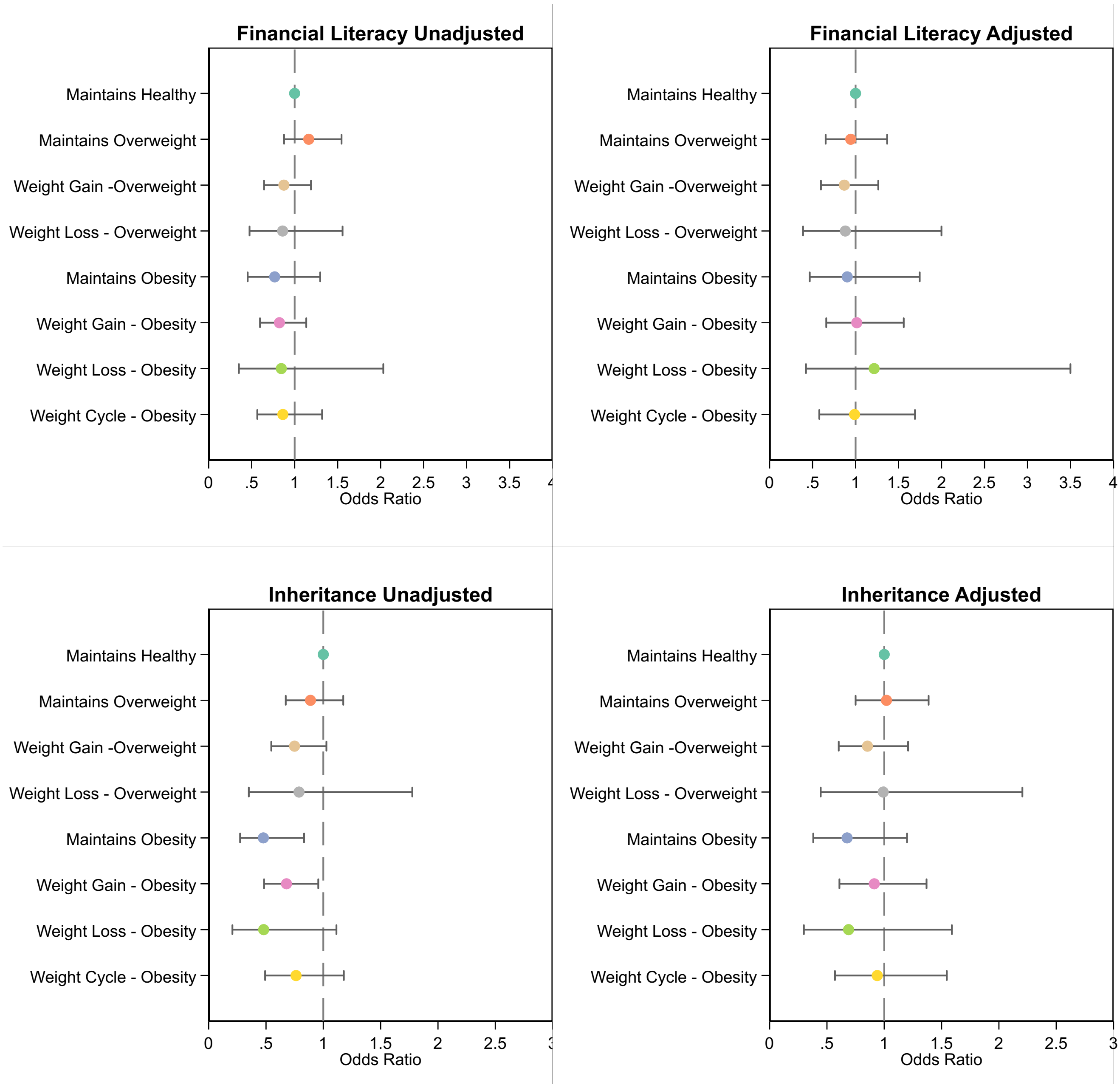

Two negative controls were used. These were selected as they would be expected to be associated with weight loss at age 50-55 only through confounding pathways:

- Financial literacy at age 62: getting three financial literacy questions correct (1: all correct; 0: at least one wrong)
- Receipt of inheritance at age 62: receiving two or more inheritances or one/none.

The tested adjustment set are the early life and age 23 confounders used in the main models.

- Sex at birth
- Age at outcome measure
- *Early life confounders:* Childhood cognition (age 11); parental social class at birth (age 0); birthweight (age 0); adolescent BMI (age 16); adolescent psychological distress (age 16).

- *Adult confounders at age 23:* self-rated health, smoking status, occupational social class, marital status, voting behaviour, trade union membership, educational attainment, income, and membership in civic or social organisations.

In both cases, adjustment for the covariate set attenuated associations between weight change group and the negative control outcome towards the null. For financial literacy, associations were fully attenuated. For receipt of inheritance, associations were reduced to non-significant.

#### S8. A note on HDL cholesterol

HDL cholesterol (HDLc) is a regular feature of blood lipid profiles and there are guidelines for what are considered "healthy levels" of HDLc in the UK (>1 mmol/L for men and >1.2 mmol/L for women) [1]. Low HDLc is associated with worse cardiovascular disease outcomes and with higher mortality in large observational studies [2]. More recent studies have however challenged the idea that the association between HDLc and cardiovascular outcomes is linear (i.e., that higher HDLc is universally protective), suggesting instead that the relationship may be U-shaped, with both low and high levels of HDLc being associated with worse outcomes [2, 3]. There is ongoing debate over the causal role of HDLc in cardiovascular disease. The biological mechanisms linking HDLc levels and cardiovascular outcomes are not yet fully understood [4]. Interventions to increase HDLc do not appear to improve cardiovascular outcomes [2], and Mendelian randomisation studies have raised questions over the role of unmeasured confounding in the negative associations between HDLc and poor health outcomes seen in observational studies [4].

[1] National Health Service. High cholesterol – Cholesterol levels [Internet]. *nhs.uk,* 2019 [cited 2025 May 14]. Available from: https://www.nhs.uk/conditions/high-cholesterol/cholesterol-levels/

[2] Perswani P, Ismail SM, Mumtaz H, et al. Rethinking HDL-C: An In-Depth NarrativeReview of Its Role in Cardiovascular Health. Curr Prob Cardiol. 2024;49(2):102152.

[3] Hamer M, O'Donovan G, Stamatakis E. High-Density Lipoprotein Cholesterol andMortality: Too Much of a Good Thing? Arterioscler Thromb Vasc Biol. American HeartAssociation; 2018;38(3):669–672.

[4] Davey Smith G, Phillips AN. Correlation without a cause: an epidemiological odyssey. *Int J Epidemiol*. 2020;49(1):4–14.
